# Elevated COVID-19 outcomes among persons living with diagnosed HIV infection in New York State: Results from a population-level match of HIV, COVID-19, and hospitalization databases

**DOI:** 10.1101/2020.11.04.20226118

**Authors:** James M. Tesoriero, Carol-Ann E. Swain, Jennifer L. Pierce, Lucila Zamboni, Meng Wu, David R. Holtgrave, Charles J. Gonzalez, Tomoko Udo, Johanne E. Morne, Rachel Hart-Malloy, Deepa T. Rajulu, Shu-Yin John Leung, Eli S. Rosenberg

**Affiliations:** New York State Department of Health, Albany, NY; Department of Health Policy Management and Behavior, University at Albany School of Public Health, State University of New York, Rensselaer, NY; Department of Epidemiology and Biostatistics, University at Albany School of Public Health, State University of New York, Rensselaer, NY; Center for Collaborative HIV Research in Practice and Policy, University at Albany School of Public Health, State University of New York, Rensselaer, NY

## Abstract

**Background:** New York State (NYS) has been an epicenter for both COVID-19 and HIV/AIDS epidemics. Persons Living with diagnosed HIV (PLWDH) may be more prone to COVID-19 infection and severe outcomes, yet few population-based studies have assessed the extent to which PLWDH are diagnosed, hospitalized, and have died with COVID-19, relative to non-PLWDH.

**Methods:** NYS HIV surveillance, COVID-19 laboratory confirmed diagnoses, and hospitalization databases were matched. COVID-19 diagnoses, hospitalization, and in-hospital death rates comparing PLWDH to non-PLWDH were computed, with unadjusted rate ratios (RR) and indirect standardized RR (sRR), adjusting for sex, age, and region. Adjusted RR (aRR) for outcomes among PLWDH were assessed by age/CD4-defined HIV disease stage, and viral load suppression, using Poisson regression models.

**Results:** From March 1-June 7, 2020, PLWDH were more frequently diagnosed with COVID-19 than non-PLWDH in unadjusted (RR [95% confidence interval (CI)]: 1.43[1.38-1.48), 2,988 PLWDH], but not in adjusted comparisons (sRR [95% CI]: 0.94[0.91-0.97]). Per-population COVID-19 hospitalization was higher among PLWDH (RR [95% CI]: 2.61[2.45-2.79], sRR [95% CI]: 1.38[1.29-1.47], 896 PLWDH), as was in-hospital death (RR [95% CI]: 2.55[2.22-2.93], sRR [95%CI]: 1.23 [1.07-1.40], 207 PLWDH), albeit not among those hospitalized (sRR [95% CI]: 0.96[0.83-1.09]). Among PLWDH, hospitalization risk increased with disease progression from HIV Stage 1 to Stage 2 (aRR [95% CI]:1.27[1.09-1.47]) and Stage 3 (aRR [95% CI]: 1.54[1.24-1.91]), and for those virally unsuppressed (aRR [95% CI]: 1.54[1.24-1.91]).

**Conclusion:** PLWDH experienced poorer COVID-related outcomes relative to non-PLWDH, with 1-in-522 PLWDH dying with COVID-19, seemingly driven by higher rates of severe disease requiring hospitalization.

## Introduction

Novel coronavirus disease 2019 (COVID-19) has resulted in the deaths of more than 1 million persons worldwide as of October 21, 2020, with the United States (US) leading the world in diagnosed cases (8,404,743) and deaths (223,000).^1^ In addition to an older, male demographic distribution that favors increased COVID-19 diagnoses, persons living with diagnosed human immunodeficiency virus (HIV) infection (PLWDH) have higher prevalence of many underlying medical conditions associated with more severe COVID-19 illness.^2-4^

The Centers for Disease Control and Prevention (CDC) currently identifies older adults and those with underlying medical conditions including chronic kidney disease, chronic obstructive pulmonary disease, obesity, sickle cell disease, diabetes, transplant recipients and those with serious heart conditions to be at elevated risk for severe illness from COVID-19. PLWDH with a low CD4 cell count or not on HIV treatment are currently listed by the CDC as *possible* risk factors for severe illness from COVID-19.^5^ Little has been firmly established regarding the extent to which PLWDH are acquiring COVID-19, the severity of COVID-19 illness experienced by PLWDH, or how these distributions compare to the non-HIV population. Emerging literature is suggesting similar or better COVID-19 clinical outcomes for PLWDH compared to non-PLWDH.^4,6-12^ The majority of these studies have been small and limited to hospitalized populations of PLWDH, factors limiting generalizability and a more complete understanding of the population-based risk of developing severe COVID-19 disease requiring hospitalization. A few recent and larger studies have found increased hospitalization and mortality outcomes for PLWDH,^13-15^ although there remain no population-based studies from US jurisdictions.

New York State (NYS) has been an epicenter for both the US COVID-19 and HIV/AIDS epidemics, placing NYS in a unique position to speak to the intersection of these two epidemics. As of October 21, 2020, NYS has 490,134 diagnosed cases and leads the US in COVID-19 deaths at 33,396.^1^ NYS, the epicenter of the HIV epidemic in the US for decades, ranked second in the number of PLWDH and first in the rate per 100,000 population at the end of 2018.^16^ Like NYS’s COVID-19 epidemic, HIV infection is concentrated in NY City (NYC), among older adults, and among racial and ethnic minorities: In 2018, 78% of PLWDH in NYS lived in NYC, 55% were over the age of 50, and 72% were either non-Hispanic Black (40%) or Hispanic (32%) persons.^17^

We conducted a population-level match of NYS’s HIV registry against its COVID-19 diagnoses and hospitalization databases, to provide the first population-level comparison of COVID-19 outcomes between the PLWDH and non-PLWDH populations within a US jurisdiction. More specifically, we compare the continuum of rates of COVID-19 diagnoses, hospitalizations, and in-hospital deaths for PLWDH in NYS to the non-PLWDH population, and determine factors associated with these outcomes among PLWDH.^18^

## Methods

### Study Population and Data Source

This is a retrospective cohort study of individuals with polymerase chain reaction (PCR) confirmed SARS-CoV-2 infection, diagnosed between March 1, 2020 and June 7, 2020 in NYS. Data were abstracted from: 1) the NYS HIV surveillance registry, which receives name-based reports for all HIV-related laboratory tests for individuals who reside or receive HIV-related care in NYS;^19-20^ 2) the NYS Electronic Clinical Laboratory Reporting System (ECLRS), an electronic system for secure and timely transmission of reportable clinical laboratory information; and 3) the State Health Information Network for NY (SHIN-NY), a public health information exchange network connecting NYS healthcare providers.

### Outcomes

The following outcomes were evaluated among individuals with and without diagnosed HIV: 1) COVID-19 diagnoses; 2) hospitalizations; and 3) in-hospital deaths. People diagnosed with COVID-19 were identified from an ECLRS file of all PCR confirmed SARS-CoV-2 infection reported to the NYS Department of Health (NYSDOH). The subset of individuals with HIV and diagnosed COVID-19 were identified by matching records from the ECLRS file of confirmed SARS-CoV-2 infection to the NYS HIV surveillance registry. Data were matched using a validated deterministic matching algorithm implemented using SAS DataFlux Data Management Studio 2.7.^21^ HIV surveillance data may require up to one year to ensure quality and completeness of information for individuals who are newly diagnosed. Therefore, PLWDH and residing in NYS as of December 2019 were included in these analyses.

COVID-19 hospitalizations are routinely identified by ongoing matching of ECLRS data to the SHIN-NY database.^22^ The SHIN-NY datafile contained hospital encounter data from January 1, 2020 to June 15, 2020 with admission dates 14 days prior to 30 days after a positive COVID-19 test result. Individuals were considered hospitalized due to COVID-19 if they had: 1) a positive COVID-19 test result ≤3 days after the discharge date; 2) an admission date up to 30 days after a positive COVID-19 test result; or 3) a positive COVID-19 test result during the hospital encounter period. An in-hospital death was defined by discharge status code of 20-21, 40-41 or 42, as defined by the HL7 v2.5 discharge disposition value set,^23^ or a discharge description indicative of patient expiration.

### Study Variables

The demographic variables included in all analyses were age, sex, and region of residence. For analyses among PLWDH, race and ethnicity, HIV transmission risk at diagnosis^24^, in HIV-related care, stage of HIV infection at last test, and viral load suppression at last test were available. CD4, viral load and genotype tests in the 3 years prior to the study period were included for analysis. In care was defined as having a CD4, viral load or genotype test reported to the HIV surveillance registry in the 365 days prior to March 1; HIV disease stage at last CD4 and viral suppression <200 copies/mL at last test were evaluated in the previous 3 years.^25,26^

### Statistical analyses

COVID-19 diagnoses, hospitalizations, and in-hospital deaths were evaluated between PLWDH and persons without diagnosed HIV (non-PLWDH).^27^ Rates per 1,000 population, and rates per previous outcome (hospitalized per diagnosis, in-hospital death per hospitalization), and unadjusted rate ratios (RR) with 95% confidence intervals (CI) were calculated to evaluate associations between study variables and each outcome. Adjusted comparisons were made via indirect standardization, controlling for age, sex, and region.^28^

Among PLWDH, rates per 1,000 population and per previous outcome, with unadjusted RRs with 95% CIs were assessed among levels of age, sex, region, race and ethnicity, transmission risk, care status, HIV stage, and viral suppression status. Adjusted RRs with 95% CI were calculated in multivariable Poisson regression models, which included covariates significant at α=0.05 in bivariate analyses. All analyses were conducted using SAS software V9.4.^21^ For these models, missing values were imputed for HIV transmission risk (12%), race and ethnicity (<1%), and age (<1%) using fully conditional specification, implemented using SAS PROC MI.^29-30^ Multiple imputation was not implemented for laboratory variables with missing data. The NYSDOH IRB approved this study.

## Results

From March 1-June 7, 2020, among 108,062 PLWDH in NYS, 2,988 were diagnosed with COVID-19 (rate: 27.65/1,000), a rate 1.43-fold (95% CI: 1.38-1.48) that among non-PLWDH (rate: 19.40/1,000) (Table 1). Similarly, elevated rates were observed across age categories, except for persons age 40-59 years, sex, and region of residence at HIV diagnosis. Standardization for these factors yielded an overall adjusted diagnosis rate-ratio (sRR) of 0.94 (95% CI: 0.91-0.97), comparing PLWDH vs. non-PLWDH (Figure 1). Standardized RR were significantly above 1.0 in regions outside of NYC, but lower in NYC (Supplementary Table 1).

**Table 1.**
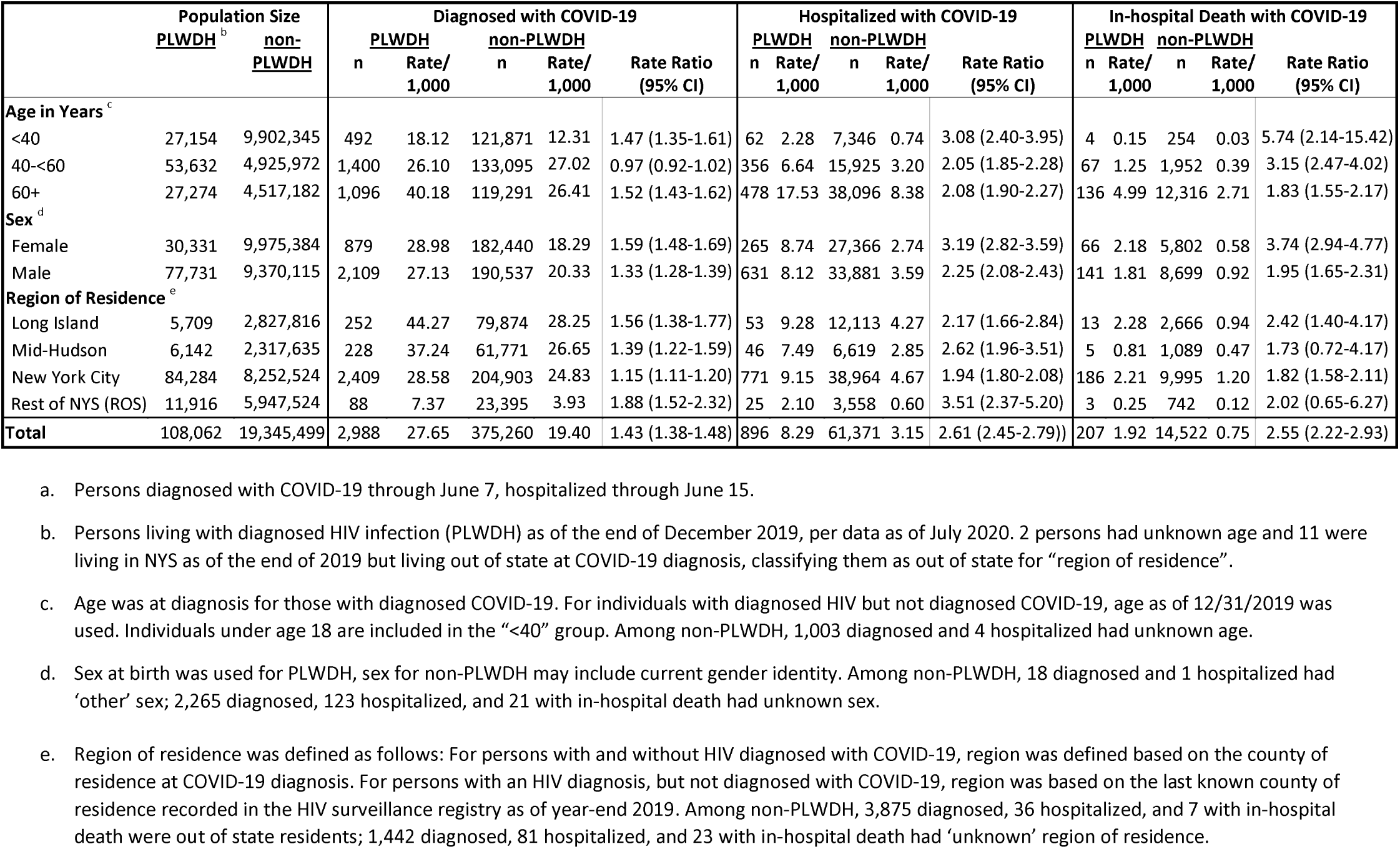
COVID-19 diagnosis, hospitalization, and in-hospital death per 1,000, among persons living with and without diagnosed HIV infection - New York State (NYS), March 1 – June 7, 2020^a^

**Figure:**
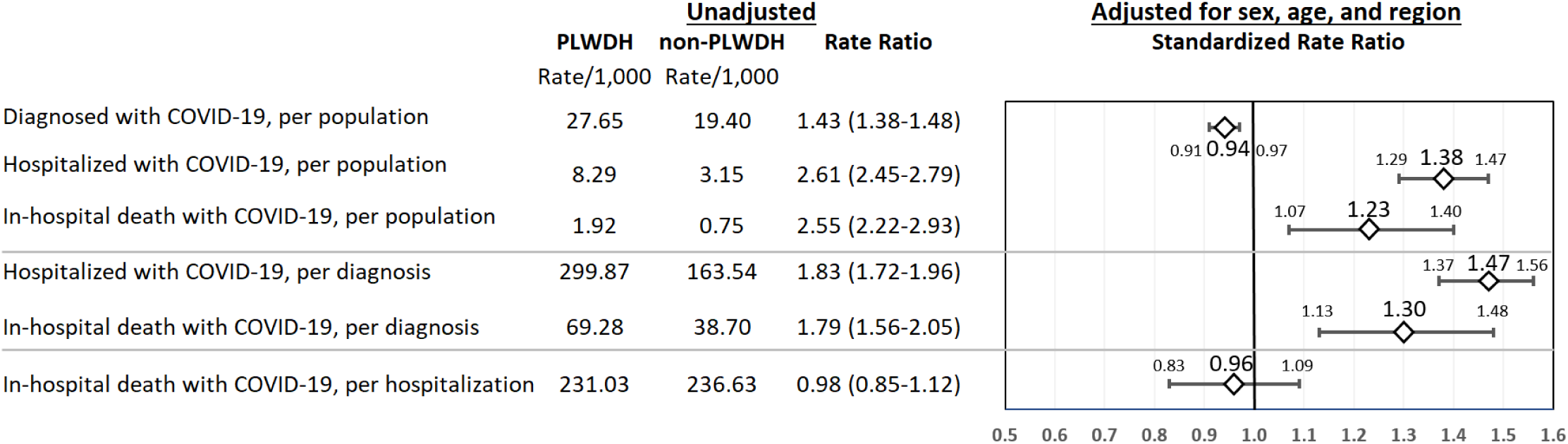
Summary of rates and rate ratios for COVID-19 diagnosis, hospitalization, and in-hospital death, comparing persons living with and without diagnosed HIV infection, by region - New York State, March 1 – June 7, 2020^a^. a. Persons diagnosed with COVID-19 through June 7, hospitalized through June 15. Standardized rate ratios adjusted for age, sex, and region.

Per-population rates of COVID-19 hospitalization were significantly elevated among PLWDH (8.29/1,000) versus non-PLWDH (3.15/1,000, RR [95% CI]: 2.61 [2.45-2.79]) and consistently so across age, sex, and geography. In unadjusted analyses, relative hospitalization for PLWDH vs. non-PLWDH was highest for persons <40 years (RR [95% CI]: 3.08 [2.40-3.95]), females (RR [95% CI]: 3.19: [2.82-3.59]), and those living in rest of NYS (ROS) (RR [95% CI]: 3.51 [2.37-5.20]). Following standardization, the disparity in hospitalization between PLWDH and non-PLWDH diminished but remained significantly elevated (sRR [95% CI]: 1.38 [1.29-1.47]).

Overall, 207 PLWDH (rate: 1.92/1,000) had a COVID-19 diagnosis and died in-hospital, at 2.55-fold (95% CI: [2.22-2.93]) the rate of the non-PLWDH population. Unadjusted per-population relative mortality was highest among persons <40 years (RR: 5.74, 95% CI: 2.14-15.42), females (3.74 (2.94-4.77), and residents of Long Island (RR [95% CI]: 2.42 [1.40-4.17]). Following adjustment, the standardized mortality ratio was 1.23 (95% CI: 1.13-1.48).

The conditional rates per previous outcomes stage for PLWDH vs. non-PLWDH are summarized in Figure 1, alongside population-level rates, and in Supplementary 2. Among those diagnosed with COVID-19, nearly one-third (299.87 per 1,000) of PLWDH were hospitalized, a rate 1.83-fold (95% CI: 1.72-1.96) that of non-PLWDH. Among those hospitalized with COVID-19, no differences were seen in in-hospital death, comparing PLWDH vs. non-PLWDH (rate-ratio: 0.98, 95% CI: [0.85-1.12], sRR: 0.96, 95% CI: [0.83-1.09]). Despite a lack of significant difference in adjusted in-hospital mortality conditional on hospitalization, the higher levels of hospitalization for PLWDH underpinned significantly higher mortality rates per-population (reported above) and per-diagnosis (case-fatality rate, 69.28/1,000 vs. 38.70/1,000, sRR: 1.30, 95% CI: [1.13-1.43]).

Among PLWDH, in bivariate analyses, COVID-19 diagnosis was associated with all factors examined, except for sex, which were included in a multivariable model (Table 2). In the adjusted model, PLWDH of older age, race and ethnicity not white non-Hispanic, and living in the regions of metropolitan NYC were significantly more likely to be diagnosed with COVID-19. No significant differences were observed between the main HIV transmission risk groups. Controlling for these factors, having Stage 3 HIV infection (aRR [95% CI] vs. Stage 1: 1.38 [1.21-1.59]) was associated with increased COVID-19 diagnosis, whereas unsuppressed viral load (aRR [95% CI] vs. suppressed: 0.70 [1.21-1.59]) was associated with decreased COVID-19 diagnosis.

**Table 2.**
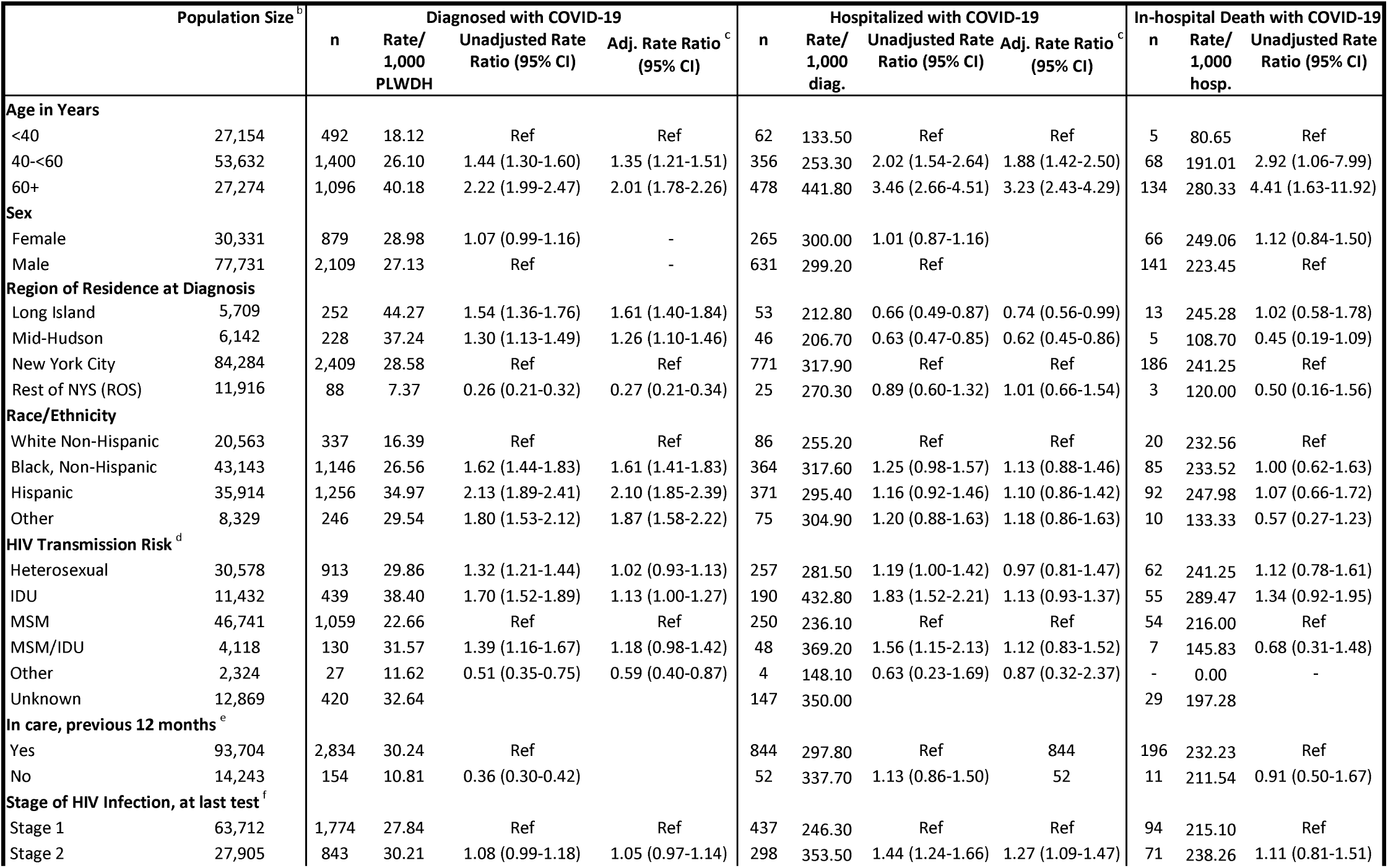
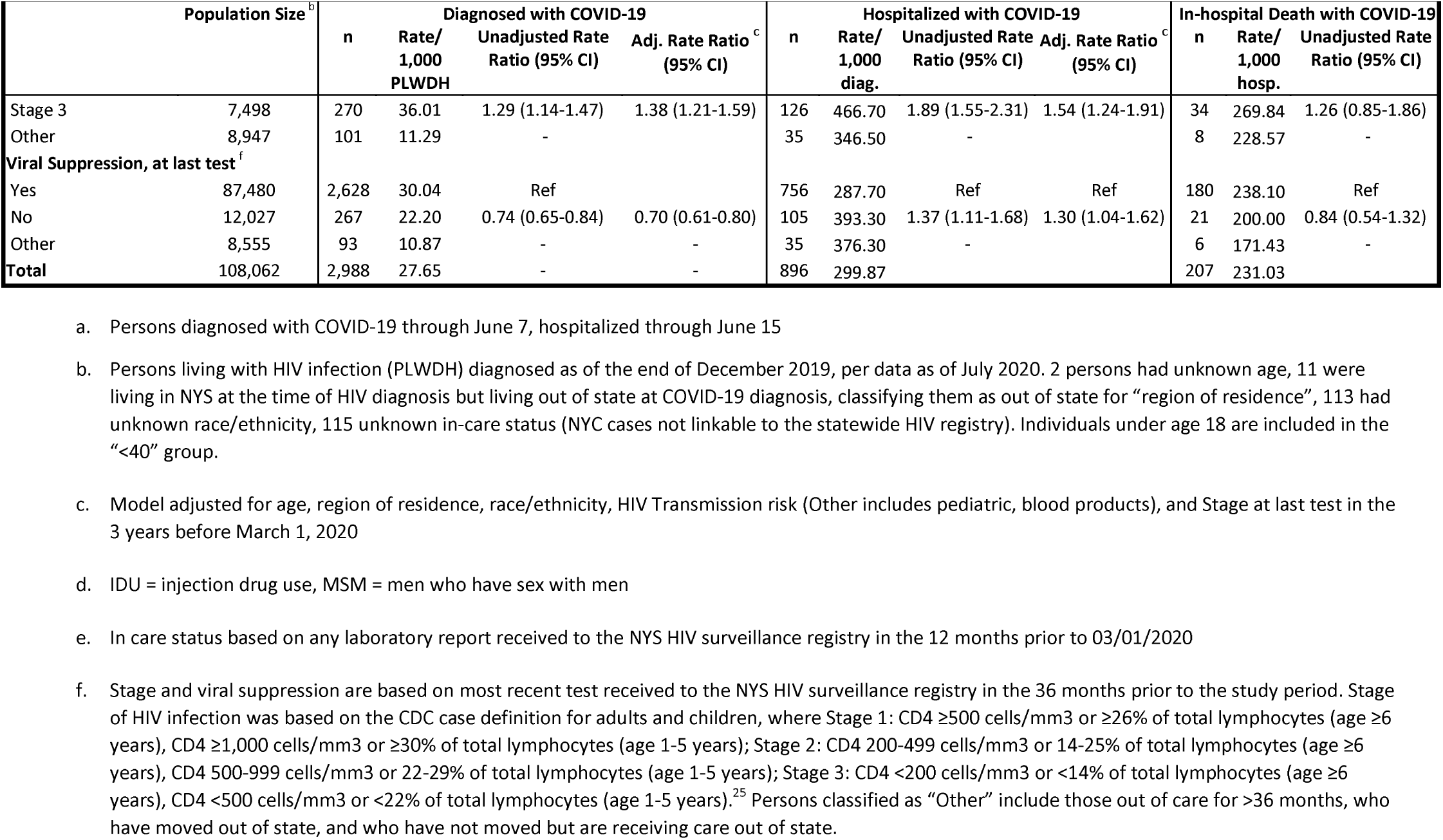
Factors associated with stages of COVID-19 diagnosis, hospitalization, and in-hospital death among persons living with diagnosed HIV infection - New York State (NYS), March 1 – June 7, 2020 ^a^

Further examining the risk-factors for hospitalization among PLWDH with diagnosed COVID-19, in adjusted analyses older age and region were associated with hospitalization, but not race, ethnicity or transmission risk (Table 3). Relative to Stage 1 infection, there was a gradient of increased hospitalization risk across Stage 2 (aRR [95% CI]: 1.27 [1.09-1.47])) and Stage 3 (aRR [95% CI]: 1.54 [1.24-1.91]). Those with unsuppressed viral load had 30% increased likelihood of hospitalization (aRR [95% CI]: 1.54 [1.24-1.91]). Among those hospitalized, only older age was associated with an increased likelihood of in-hospital death.

To probe the role of HIV stage and viral suppression in increasing hospitalization risk for PLWDH versus non-PLWDH, we conducted the per-population hospitalization standardized rate analysis classifying PLWDH persons into 2 categories: PLWDH Stage 1 and virally suppressed, PLWDH in other known care statuses. Relative to non-PLWDH, hospitalization risk was elevated even for PLWDH Stage 1 and virally suppressed (sRR [95% CI]: 1.19 [1.07-1.30]), along with PLWDH in other care statuses (sRR [95% CI]: 1.77 [1.61-1.93]).

## Discussion

Our study represents the first population-level match of a US state’s HIV registry against its COVID-19 diagnoses and hospitalization databases, establishing state-level rates of COVID-19 outcomes among the PLWDH population, and state-level comparisons of these rates to those observed in the overall population.

### COVID-19 Diagnosis

2.8% of NYS’s PLWDH population had been diagnosed with COVID through June 7, 2020, a rate nearly 40% higher than that observed in the non-PLWDH population. This disparity disappeared after standardization, a finding consistent with a meta-analysis of 14 smaller studies (8 from the US) finding a higher but not statistically different rate of COVID-19 diagnoses among PLWDH.^31^ After adjusting for age, region of residence, race and ethnicity, HIV transmission risk, and HIV stage at last test, COVID-19 diagnoses rates among PLWDH did not differ by sex at birth or transmission risk. Diagnosis rates were, significantly higher among PLWDH age 40 and over, a finding reported elsewhere.^32^ Consistent with a convenience sample of PLWDH,^2^ our study found a higher-than-background rate of COVID-19 in PLWDH of color: Adjusted diagnosis rates were 1.6 times higher for non-Hispanic Blacks and 2.1 times higher for Hispanics compared to non-Hispanic Whites. This finding parallels the racial and ethnic differences in the distribution of HIV in NYS,^17^ and may reflect the impact of differential rates of COVID-19-enhancing comorbidities among PLWDH of color, and/or social and behavioral determinants of health associated with COVID-19 transmission in minority communities.^33-36^

COVID-19 diagnosis rates among PLWDH also varied by region. Consistent with overall population rates, we found significantly lower diagnoses rates among PLWDH in upstate New York (ROS). Diagnoses rates among PLWDH were significantly higher in the NYC-adjacent regions of Long Island and Mid-Hudson than they were in NYC. This finding may reflect lower COVID-19 testing availability in NYC during the initial phase of the pandemic rather than a difference in background infection levels. Although there were no overall differences in COVID-19 diagnoses between PLWDH and non-PLWDH, PLWDH exhibited higher rates outside NYC and a lower rate within NYC. Because we were not able to standardize the comparative analyses by race and ethnicity, this finding may reflect differences in the racial and ethnic distribution of PLWDH relative to the overall population by region, rather than a substantive difference in diagnosis propensity among PLWDH.

The relationship between the management of HIV infection and COVID-19 diagnosis was not straightforward. Virally suppressed PLWDH were significantly *more* likely to have been diagnosed, as were PLWDH with CD4 counts below 200 cell/mm^3^. The increased diagnosis probability among virally suppressed persons may reflect a difference in test-seeking behavior or more interaction with the health care system among suppressed PLWDH rather than a difference in underlying COVID-19 prevalence. Alternatively, healthier PLWDH may place themselves at greater risk for acquiring COVID-19, although this interpretation is inconsistent with findings from one existing study.^3^

### COVID-19 Hospitalization and Mortality

PLWDH were significantly more likely than non-PLWDH to be hospitalized with COVID-19, overall (sRR=1.38) and among diagnosed cases (sRR=1.47), suggesting higher rates of severe disease among PLWDH requiring hospitalization. Hospitalization rates among PLWDH were higher among those not virally suppressed and those with lower CD4 counts, suggesting that less controlled HIV virus may increase COVID-19 severity to where hospitalization is required. Supplemental data analyses found higher hospitalization rates compared to the non-PLWDH population persisted among the subset of PLWDH who were virally suppressed with high CD4 counts, suggesting that additional factors may explain elevated hospitalization rates among PLWDH, including other comorbidities, systemic stress of chronic viral infection, and social determinants of COVID-19 severity. Hospitalization rates were also higher among those over 40 and those with a documented injection drug use history, the latter finding also observed in a non-HIV-focused case-control study spanning 50 US states.^37^

We observed elevated population-level mortality for PLWDH which was driven by higher hospitalization rates and not higher mortality among hospitalized PLWDH. The unadjusted case fatality rate among PLWDH was nearly twice that exhibited in the non-PLWDH population, with a significant difference maintained but attenuated after statistical adjustment. The number of COVID-19 deaths among PLWDH constitutes a sizable increase over normal levels. There were 490 deaths among PLWDH from March 1-June 15, 2019. Against this backdrop, the 207 COVID-19-specific hospital deaths in our study represents a 42% addition to anticipated deaths during this period. Further analyses refining this estimate are needed.^38^ Higher mortality among PLWDH was reported in a large population cohort study of health care facility attendees in Western Cape of South Africa,^13^ in cohorts of hospitalized patients in London^14^**Error! Bookmark not defined**. and NYC,^15^ and in a study of PLWDH receiving antiretroviral therapy in 60 Spanish hospitals.^12^ Contrary findings have been reported in a NYC study comparing 88 COVID-19 hospitalized PLWDH to a matched non-PLWDH control group.^4^

The only statistically significant predictor of in-hospital mortality among hospitalized PLWDH was age, with those age 40 and older 3-4 times more likely to experience in-hospital death depending on age group. Given the well-established findings on elevated mortality by increasing age regardless of HIV-status^13,32,39^ this likely reflects an elevated risk of COVID-19 severity-enhancing comorbidities among older adults, including diabetes, hypertension, and chronic lung and cardiovascular disease. Also reported in the literature,^12,40^we found that hospitalized and fatal COVID-19 cases were younger among PLWDH. This finding may lend support to the notion that HIV infection can accelerate biological aging.^41,42^

Although non-Hispanic Black and Hispanic PLWDH were more likely to be diagnosed with COVID-19 than non-Hispanic white PLWDH, they were not more likely to be hospitalized once diagnosed, or to die once hospitalized. This finding is partially consistent with COVID-19 studies finding racial and ethnic disparity present in hospitalization rates but not in mortality.^18,43-45^ Finally, despite RRs in the expected direction, CD4 count was not significantly related to in-hospital death. This finding is incongruent with at least 2 studies finding that CD4 counts less than 200 cell/mm^3^ were significant predictors of decreased survival probability among hospitalized PLWDH.^13,32^

### Limitations

This earliest outcome in this study is COVID-19 laboratory confirmed diagnosis and not infection. A statewide seroprevalence study estimated that about 9% of COVID-19 cases through March 2020 had been diagnosed in NYS.^46^ Differences in diagnosis propensity among PLWDH or between PLWDH and the non-PLWDH population could alter the interpretation of some findings. Our analyses were limited to the demographic and laboratory data available in NYS’s HIV surveillance and COVID-19 registries, precluding more in-depth investigations into the role played by co-morbidities and underlying medical conditions, COVID-19 risk behaviors, and social determinants of health. It is important to further investigate how the observed associations may be changed by information on co-morbidities and underlying medical conditions, COVID-19 risk behaviors, and social determinants of health with more comprehensive data sources such as medical chart reviews. Our denominator of PLWDH included people who died between January 1 and June 15, 2020 and excluded persons newly diagnosed with HIV during this same timeframe. Because these numbers have historically offset each other, the impact of this limitation is likely negligible.

## Conclusion

Controlling for age, sex, and region, PLWDH in NYS were diagnosed with COVID-19 at roughly the same rate as non-PLWDH. Further, once hospitalized, PLWDH had similar mortality rates compared to non-PLWDH. However, PLWDH were hospitalized at a 40% higher rate, with low CD4 and high VL linked to higher hospitalization, and ultimately deaths, among PLWDH, although persons with well-managed HIV per CD4 and viral load still had elevated hospitalization rates. This surveillance approach has illuminated increased COVID-19 morbidity and mortality for PLWDH, while highlighting the need to further understand the mechanisms underpinning this increased risk of severe disease.

## Supporting information

Supplemental matrial

## Data Availability

The public health surveillance data reported on in this manuscript are not publicly available

## Acknowledgement

The co-authors wish to acknowledge the following essential contributors to this work. Amy Kelly for literature review and editing contributions. Dr. Heather Bradley, Georgia State University, for input on the organization of findings. We acknowledge funding from NIH 1R01DA051302 (ESR).

## References

1. Johns Hopkins University. COVID-19 United States Cases 2020. https://coronavirus.jhu.edu/us-map.

2. Meyerowitz EA, Kim AY, Ard KL, et al. Disproportionate burden of COVID-19 among racial minorities and those in congregate settings among a large cohort of people with HIV. AIDS. 2020;34(12):1781–1787. doi: 10.1097/QAD.0000000000002607.

3. Kalichman SC, Eaton LA, Berman M, et al. Intersecting pandemics: Impact of SARS-CoV-2 (COVID-19) protective behaviors on people living with HIV, Atlanta, Georgia. J Acquir Immune Defic Syndr. 2020; 85(1):66–72. doi: 10.1097/QAI.0000000000002414.

4. Sigel K, Swartz T, Golden E, et al. Covid-19 and people with HIV infection: Outcomes for hospitalized patients in New York City [Published online June 28, 2020]. Clin Infect Dis. doi: 10.1093/cid/ciaa880.

5. Centers for Disease Control and Prevention. People of any age with underlying medical conditions. Updated June 25, 2020. https://www.cdc.gov/coronavirus/2019-ncov/need-extra-precautions/people-with-medical-conditions.html. Accessed July 13, 2020.

6. Okoh AK, Bishburg E, Grinberg S, et al. COVID-19 pneumonia in patients with HIV - A Case Series. J Acquir Immune Def Syndr. 2020;85(1):e4–e5. doi: 10.1097/QAI.0000000000002411.

7. Shalev N, Scherer M, LaSota ED, et al. Clinical characteristics and outcomes in people living with HIV hospitalized for COVID-19 [Published online May 30, 2020]. Clin Infect Dis. doi: 10.1093/cid/ciaa635.

8. Vizcarra P, Perez-Elias MJ, Quereda C, et al. Description of COVID-19 in HIV-infected individuals: a single-centre, prospective cohort. Lancet HIV. 2020;7(8):e554–e564. doi: 10.1016/S2352-3018(20)30164-8.

9. Patel VV, Felsen UR, Fisher M, et al. Clinical outcomes by HIV serostatus, CD4 count, and viral suppression among people hospitalized with COVID-19 in the Bronx, New York. Presented at 23rd International AIDS Conference, 2020.

10. Guo W, Ming, F, Dong Y, et al. A Survey for COVID-19 among HIV/AIDS Patients in Two Districts of Wuhan, China (3/4/2020). http://dx.doi.org/10.2139/ssrn.3550029.

11. Karmen-Tuohy S, Carlucci PM, Zervou FN, et al. Outcomes among HIV-positive patients hospitalized with COVID-19. J Acquir Immune Defic Syndr. 2020;85(1):6–10. doi: 10.1097/QAI.0000000000002423.

12. Gervasoni C, Meraviglia P, Riva A, et al. Clinical features and outcomes of HIV patients with coronavirus disease 2019 [Published on May 14, 2020]. Clin Infect Dis. doi.: 10.1093/cid/ciaa579.

13. Davies MA. HIV and risk of COVID-19 death: a population cohort study from the Western Cape Province, South Africa. medRxiv. 2020. doi: 10.1101/2020.07.02.20145185.

14. Childs K, Post FA, Norcross C, et al. Hospitalized patients with COVID-19 and HIV: a case series [Published online May 27, 2020]. Clin Infect Dis. doi: 10.1093/cid/ciaa657.

15. Suwanwongse K, Shabarek N. Clinical features and outcome of HIV/SARS-CoV-2 co-infected patients in the Bronx, New York City [Published online May 28, 2020]. J Med Virol. DOI: 10.1002/jmv.26077.

16. Centers for Disease Control and Prevention. HIV Surveillance Report, 2018 (Updated); 31. http://www.cdc.gov/hiv/library/reports/hiv-surveillance.html. Published May 2020.

17. New York State Department of Health, AIDS Institute, Bureau of HIV/AIDS Epidemiology. New York State HIV/AIDS annual surveillance report for persons diagnosed through December 2018. December 2019. https://www.health.ny.gov/diseases/aids/general/statistics/annual/2018/2018_annual_surveillance_report.pdf. Accessed July 17, 2020.

18. Holtgrave DR, Barranco MA, Tesoriero JM, Blog DS, Rosenberg ES. Assessing racial and ethnic disparities using a COVID-19 outcomes continuum for New York State. Ann of Epidemiol. 2020;48:9–14. doi: 10.1016/j.annepidem.2020.06.010.

19. New York State Department of Health. Part 63 HIV/AIDS Testing, Reporting and Confidentiality of HIV-Related Information. http://www.health.ny.gov/nysdoh/rfa/hiv/full63.htm. Accessed 8/18/2020.

20. New York State. Statutory Authority: Public Health Law. Sect. §2786 and Article 21.

21. SAS Institute Inc. SAS. https://www.sas.com/en_us/home.html. Assessed 10/15/20.

22. Rosenberg ES, Dufort EM, Udo T, et al. Association of Treatment With Hydroxychloroquine or Azithromycin With In- Hospital Mortality in Patients With COVID-19 in New York State. JAMA. 2020; 323(24):2493–2502. doi: 10.1001/jama.2020.8630.

23. Caristix. HL7 v2.5 - 0112 - Discharge Disposition. 2020. https://hl7-definition.caristix.com/v2/HL7v2.5/Tables/0112. Accessed 10/15/2020.

24. Centers for Disease Control and Prevention. eHARS v4.10 Technical Reference Guide. 2018.

25. Sabharwal CJ, Braunstein SL, Robbins RS, Shepard CW. Optimizing the use of surveillance data for monitoring the care status of persons recently diagnosed with HIV in NYC. J Acquir Immune Defic Syndr. 2014;65(5):571–578. doi: 10.1097/QAI.0000000000000077.

26. Swain CA, Smith LC, Nash D, et al. Postpartum Human Immunodeficiency Virus Care Among Women Diagnosed During Pregnancy. Obstet Gynecol. 2016;128(1):44–51. doi: 10.1097/AOG.0000000000001454.

27. National Center for Health Statistics. Estimates of the April 1, 2010 resident population of the United States, by county, single-year of age, bridged race, Hispanic origin, and sex. https://www.cdc.gov/nchs/nvss/bridged_race.htm.

28. North Carolina Public Health. Age-adjusted Death Rates. In: Statistics SCfH, ed. Raleigh, NC 27699- 19082010.

29. van Buuren S. Multiple imputation of discrete and continuous data by fully conditional specification. Stat Methods Med Res. 2007;16(3):219-242. doi 10.1177/0962280206074463.

30. SAS Institute Inc. SAS/STAT® 14.1 User’s Guide, The MI Procedure. https://support.sas.com/documentation/onlinedoc/stat/141/mi.pdf. Accessed 10/15/2020.

31. Ssentongo P, Heilbrunnes ES, Ssentongo AE, et al. Prevalence of HIV in patients hospitalized for COVID-19 and associated outcomes: a systematic review and meta-analysis. medRxiv. 2020. doi: 10.1101/2020.07.03.20143628.

32. Del Amo J, Polo R, Moreno S, et al. Incidence and severity of COVID-19 in HIV positive persons receiving antiretroviral therapy. A cohort study. Ann Intern Med. 2020;173(7):536–541. doi: 10.7326/M20-3689.

33. Huang VS, Sutermaster S, Caplan Y, et al. Social distancing across vulnerability, race, politics, and employment: How different Americans changed behaviors before and after major COVID-19 policy announcements. medRxiv. 2020. doi: 10.1101/2020.06.04.20119131.

34. Millett GA, Honermann B, Jones A, et al. White Counties Stand Apart: The Primacy of Residential Segregation in COVID-19 and HIV Diagnoses. AIDS Patient Care and STDs. 2020;34(10):417–424. doi: 10.1089/apc.2020.0155.

35. Millett GA, Jones AT, Benkeser D, et al. Assessing differential impacts of COVID-19 on black communities. Ann Epidemiol. 2020;47:37–44. doi: 10.1016/j.annepidem.2020.05.003.

36. Rodriguez-Diaz CE, Guilamo-Ramos V, Mena L, et al. Risk for COVID-19 infection and death among Latinos in the United States: Examining heterogeneity in transmission dynamics [Published online July 23, 2020]. Ann Epidemiol. doi: 10.1016/j.annepidem.2020.07.007.

37. Wang QQ, Kaelber DC, Xu R. et al. COVID-19 risk and outcomes in patients with substance use disorders: analyses from electronic health records in the United States [Published online September 30, 2020]. Mol Psychiatry. doi: 10.1038/s41380-020-00895-0.

38. Rossen LM, Branum AM, Ahmad FB, et al. Excess Deaths Associated with COVID-19, by Age and Race and Ethnicity - United States, January 26-October 3, 2020. MMWR. 2020;69(42):1522–1527. doi: 10.15585/mmwr.mm6942e2.

39. Dandachi D, Geiger G, Montgomery MW, et al. Characteristics, Comorbidities, and Outcomes in a Multicenter Registry of Patients with HIV and Coronavirus Disease-19 [Published online September 9, 2020]. Clin Infect Dis. doi: 10.1093/cid/ciaa1339.

40. Blanco JL, Ambrosioni J, Garcia F, et al. COVID-19 in patients with HIV: clinical case series. Lancet HIV. 2020;7(5):e314–e316. doi: 10.1016/S2352-3018(20)30111-9.

41. De Francesco D, Wit FW, Bürkle A, et al. Do people living with HIV experience greater age advancement than their HIV-negative counterparts? AIDS. 2019;33(2):259–268. doi: 10.1097/QAD.0000000000002063.

42. Lagathu C, Cossarizza A, Béréziat V, et al. Basic science and pathogenesis of ageing with HIV: potential mechanisms and biomarkers. AIDS. 2017;31(Suppl 2):S105–S119. doi: 10.1097/QAD.0000000000001441.

43. Azar KMJ, Shen Z, Romanelli RJ, Lockhart SH, et al. Disparities In Outcomes Among COVID-19 Patients In A Large Health Care System In California. Health Aff (Millwood). 2020;39(7):1253–1262. doi: 10.1377/hlthaff.2020.00598.

44. Rentsch CT, Kidwai-Khan F, Tate JP, et al. Covid-19 by Race and Ethnicity: A National Cohort Study of 6 Million United States Veterans. medRxiv. 2020. doi: 10.1101/2020.05.12.20099135.

45. Price-Haywood EG, Burton J, Fort D, Seoane L. Hospitalization and Mortality among Black Patients and White Patients with Covid-19. N Eng J Med. 2020;382(26):2534–2543. doi: 10.1056/NEJMsa2011686.

46. Rosenberg ES, Tesoriero JM, Rosenthal EM, et al. Cumulative incidence and diagnosis of SARS-CoV-2 infection in New York. Ann Epidemiol. 2020;48:23-29.e4. doi: 10.1016/j.annepidem.2020.06.004.

