## Supplemental matrial for "Elevated COVID-19 outcomes among persons living with diagnosed HIV infection in New York State: Results from a population-level match of HIV, COVID-19, and hospitalization databases"

**Supplementary Material**

James M. Tesoriero PhD^a,b,d^, Carol-Ann E. Swain PhD^a^, Jennifer L. Pierce BS^a^, Lucila Zamboni PhD^a^, Meng Wu PhD^a^, David R. Holtgrave PhD^b,d^, Charles J. Gonzalez MD^a^, Tomoko Udo PhD^b,d^, Johanne E. Morne MS^a,d^, Rachel Hart-Malloy PhD^a,c,d^, Deepa T. Rajulu MS^a^, Shu-Yin John Leung MA^a^, Eli S. Rosenberg PhD^c,d^

a. New York State Department of Health, Albany, NY

b. Department of Health Policy Management and Behavior, University at Albany School of Public Health, State University of New York, Rensselaer, NY

c. Department of Epidemiology and Biostatistics, University at Albany School of Public Health, State University of New York, Rensselaer, NY

d. Center for Collaborative HIV Research in Practice and Policy, University at Albany School of Public Health, State University of New York, Rensselaer, NY

**Supplementary Table 1.** Age- and sex- standardized rate ratios for COVID-19 diagnosis, hospitalization, and in-hospital death, comparing persons living with and without diagnosed HIV infection, by region - New York State, March 1 – June 7, 2020 ^a^

|  | **Diagnosed with COVID-19** | **Hospitalized with COVID-19** | **In-hospital Death with COVID-19** |
| --- | --- | --- | --- |
|  | **Standardized Rate Ratio  (95% CI)** | **Standardized Rate Ratio  (95% CI)** | **Standardized Rate Ratio  (95% CI)** |
| **Rates per 1,000 population** |  |  |  |
| Long Island | 1.33 (1.17-1.50) | 1.64 (1.20-2.08) | 1.80 (0.82-2.79) |
| Mid-Hudson | 1.14 (1.00-1.30) | 1.89 (1.34-2.43) | 1.23 (0.15-2.31) |
| New York City | 0.88 (0.85-0.92) | 1.32 (1.23-1.41) | 1.20 (1.03-1.37) |
| Rest of New York State (ROS) | 1.81 (1.43-2.19) | 2.91 (1.77-4.06) | 1.85 (0.00-3.95) |
| **Rates per previous stage** ^b^ |  |  |  |
| Long Island | 1.34 (1.17-1.50) | 1.27 (0.93-1.62) | 1.09 (0.50-1.68) |
| Mid-Hudson | 1.15 (1.00-1.30) | 1.50 (1.07-1.94) | 0.68 (0.08-1.28) |
| New York City | 0.90 (0.86-0.93) | 1.47 (1.37-1.58) | 0.96 (0.83-1.10) |
| Rest of New York State (ROS) | 1.81 (1.44-2.19) | 1.80 (1.09-2.50) | 0.81 (0.00-1.72) |

a. Persons diagnosed with COVID-19 through June 7, hospitalized through June 15
b. Denominator for “Diagnosed with COVID-19” is population, for “Hospitalized with COVID-19” is persons diagnosed with COVID-19, for “In-hospital Death with COVID-19” is persons hospitalized with COVID-19.

**Supplementary Table 2.** COVID-19 diagnosis, hospitalization, and in-hospital death per 1,000, among persons living with and without diagnosed HIV infection - New York State (NYS), March 1 – June 7, 2020 ^a^

|  | **Population Size** | | | **Diagnosed with COVID-19** | | | | | | | **Hospitalized with COVID-19** | | | | | | | **In-hospital Death with COVID-19** | | | | | | | |
| --- | --- | --- | --- | --- | --- | --- | --- | --- | --- | --- | --- | --- | --- | --- | --- | --- | --- | --- | --- | --- | --- | --- | --- | --- | --- |
|  | **PLWDH** ^b^ | **non-PLWDH** | | **PLWDH** | | | | **non-PLWDH** | |  | **PLWDH** | | **non-PLWDH** | | | |  | **PLWDH** | | | **non-PLWDH** | | | |  |
|  |  |  |  | **n** | **Rate/ 1,000** | | | **n** | **Rate/ 1,000** | **Rate Ratio  (95% CI)** | **n** | **Rate/ 1,000 diag.** | **n** | | **Rate/ 1,000 diag.** | | **Rate Ratio  (95% CI)** | **n** | **Rate/ 1,000 hosp.** | | **n** | | **Rate/ 1,000 hosp.** | | **Rate Ratio  (95% CI)** |
| **Age in Years** ^c^ |  |  | |  |  | | |  |  |  |  |  |  | |  | |  |  |  | |  | |  | |  |
| <40 | 27,154 | 9,902,345 | | 492 | 18.12 | | | 121,871 | 12.31 | 1.47 (1.35-1.61) | 62 | 126.02 | 7,346 | | 60.28 | | 2.09 (1.63-2.68) | 4 | 64.52 | | 254 | | 34.58 | | 1.87 (0.69-5.01) |
| 40-<60 | 53,632 | 4,925,972 | | 1,400 | 26.10 | | | 133,095 | 27.02 | 0.97 (0.92-1.02) | 356 | 254.29 | 15,925 | | 119.65 | | 2.13 (1.91-2.36) | 67 | 188.20 | | 1,952 | | 122.57 | | 1.54 (1.20-1.96) |
| 60+ | 27,274 | 4,517,182 | | 1,096 | 40.18 | | | 119,291 | 26.41 | 1.52 (1.43-1.62) | 478 | 436.13 | 38,096 | | 319.35 | | 1.37 (1.25-1.50) | 136 | 284.52 | | 12,316 | | 323.29 | | 0.88 (0.74-1.04) |
| **Sex** ^d^ |  |  | |  |  | | |  |  |  |  |  |  | |  | |  |  |  | |  | |  | |  |
| Female | 30,331 | 9,975,384 | | 879 | 28.98 | | | 182,440 | 18.29 | 1.59 (1.48-1.69) | 265 | 301.48 | 27,366 | | 150.00 | | 2.01 (1.78-2.27) | 66 | 249.06 | | 5,802 | | 212.01 | | 1.18 (0.92-1.50) |
| Male | 77,731 | 9,370,115 | | 2,109 | 27.13 | | | 190,537 | 20.33 | 1.33 (1.28-1.39) | 631 | 299.19 | 33,881 | | 177.82 | | 1.68 (1.56-1.82) | 141 | 223.45 | | 8,699 | | 256.75 | | 0.87 (0.74-1.03) |
| **Region of Residence** ^e^ | | |  | | |  |  | |  |  |  |  |  |  | |  | |  | |  | |  | |  | |
| Long Island | 5,709 | 2,827,816 | | 252 | 44.14 | | | 79,874 | 28.25 | 1.56 (1.38-1.77) | 53 | 210.32 | 12,113 | | 151.65 | | 1.39 (1.06-1.82) | 13 | 245.28 | | 2,666 | | 220.09 | | 1.11 (0.65-1.92) |
| Mid-Hudson | 6,142 | 2,317,635 | | 228 | 37.12 | | | 61,771 | 26.65 | 1.39 (1.22-1.59) | 46 | 201.75 | 6,619 | | 107.15 | | 1.88 (1.41-2.52) | 5 | 108.70 | | 1,089 | | 164.53 | | 0.67 (0.27-1.59) |
| New York City | 84,284 | 8,252,524 | | 2,409 | 28.58 | | | 204,903 | 24.83 | 1.15 (1.11-1.20) | 771 | 320.05 | 38,964 | | 190.16 | | 1.68 (1.57-1.81) | 186 | 241.25 | | 9,995 | | 256.52 | | 0.94 (0.81-1.09) |
| Rest of NYS | 11,916 | 5,947,524 | | 88 | 7.39 | | | 23,395 | 3.93 | 1.88 (1.52-2.32) | 25 | 284.09 | 3,558 | | 152.08 | | 1.87 (1.26-2.77) | 3 | 120.00 | | 742 | | 208.54 | | 0.58 (0.19-1.79) |
| **Total** | 108,062 | 19,345,499 | | 2,988 | 27.65 | | | 375,260 | 19.40 | 1.43 (1.38-1.48) | 896 | 299.87 | 61,371 | | 163.54 | | 1.83 (1.72-1.96) | 207 | 231.03 | | 14,522 | | 236.63 | | 0.98 (0.85-1.12) |

a. Persons diagnosed with COVID-19 through June 7, hospitalized through June 15.
b. Persons living with diagnosed HIV infection (PLWDH) as of the end of December 2019, per data as of July 2020. 2 persons had unknown age and 11 were living in NYS as of the end of 2019 but living out of state at COVID-19 diagnosis, classifying them as out of state for “region of residence”.
c. Age was at diagnosis for those with diagnosed COVID-19. For individuals with diagnosed HIV but not diagnosed COVID-19, age as of 12/31/2019 was used. Individuals under age 18 are included in the “<40” group. Among non-PLWDH, 1,003 diagnosed and 4 hospitalized had unknown age.
d. Sex at birth was used for PLWDH, sex for non-PLWDH may include current gender identity. Among non-PLWDH, 18 diagnosed and 1 hospitalized had ‘other’ sex; 2,265 diagnosed, 123 hospitalized, and 21 with in-hospital death had unknown sex.
e. Region of residence was defined as follows: For persons with and without HIV diagnosed with COVID-19, region was defined based on the county of residence at COVID-19 diagnosis. For persons with an HIV diagnosis, but not diagnosed with COVID-19, region was based on the last known county of residence recorded in the HIV surveillance registry as of year-end 2019. Among non-PLWDH, 3,875 diagnosed, 36 hospitalized, and 7 with in-hospital death were out of state residents; 1,442 diagnosed, 81 hospitalized, and 23 with in-hospital death had ‘unknown’ region of residence.
